# Comprehensive assessment of *PINK1* variants in Parkinson’s disease

**DOI:** 10.1101/2020.01.21.20018101

**Authors:** Lynne Krohn, Francis P. Grenn, Mary B. Makarious, Jonggeol Jeffrey Kim, Sara Bandres-Ciga, Dorien A. Roosen, Ziv Gan-Or, Mike A. Nalls, Andrew B. Singleton, Cornelis Blauwendraat, on behalf of the International Parkinson’s Disease Genomics Consortium (IPDGC)

## Abstract

Multiple genes have been associated with monogenic Parkinson’s disease and Parkinsonism syndromes. Mutations in *PINK1* (PARK6) have been shown to result in autosomal recessive early onset Parkinson’s disease. In the past decade, several studies have suggested that carrying a single heterozygous *PINK1* mutation is associated with increased risk for Parkinson’s disease. Here we comprehensively assess the role of *PINK1* variants in Parkinson’s disease susceptibility using several large datasets totalling 376,558 individuals including: 13,708 Parkinson’s disease cases and 362,850 controls. After combining these data, we did not find evidence to support a role for heterozygous *PINK1* mutations as a risk factor for Parkinson’s disease.

## Introduction

Parkinson’s disease (PD) is a complex disorder caused by polygenic and environmental factors. In approximately 1-2% of the (mainly familial) PD cases, a single gene is responsible for disease development. Multiple genes have been reported to cause PD or Parkinsonism either via autosomal dominant or autosomal recessive inheritance patterns (Blauwendraat, Nalls, and Singleton 2019). In 2004, mutations in *PINK1* (*PARK6*) were identified to cause PD (Valente et al. 2004). Currently, homozygous or compound heterozygous mutations in *PINK1* that result in loss-of-function are a well-known cause of early onset PD (Kasten et al. 2018). Interestingly, some studies have also suggested a potential role for heterozygous *PINK1* mutations on parkinsonism in offspring of biallelic mutation carriers (Criscuolo et al. 2006; Klein et al. 2007; Eggers et al. 2010; Ricciardi et al. 2014) or PD in potential families with a seemingly autosomal dominant pattern of inheritance for *PINK1*, in particular for *PINK1* p.G411S (rs45478900) (Abou-Sleiman et al. 2006; Toft et al. 2007; Mellick et al. 2009). This potential effect is supported by functional evidence showing that the *PINK1* p.G411S mutation interferes with the protective functions of PINK1-mediated mitochondrial quality control. Additionally, *PINK1* p.G411S heterozygous carriers seem to have a significant reduction in kinase activity (Puschmann et al. 2017). However, there is a lack of replication in large datasets showing a robust genetic association for heterozygous *PINK1* variants and PD risk. Here, we scrutinize the effect of heterozygous *PINK1* variants, including p.G411S, in a large series of PD patients including 13,708 of PD cases and 362,850 of controls of European ancestry from multiple independent datasets.

## Methods

### Parkinson’s disease and control genetic data

#### Genotyping data

Genotyping data (all Illumina platform based) was obtained from International Parkinson’s Disease Genomics Consortium (IPDGC) members, collaborators and public resources. All datasets underwent quality control separately, both on individual level data and variant level data before imputation as previously described (Nalls et al. 2019; Blauwendraat et al. 2019). NeuroX (Nalls et al. 2015) datasets (PPMI, HBS, PDBP, NeuroX_DBGAP) had directly genotyped the *PINK1* p.G411S variant with probes exm27138 and exm27138_ver2. See Supplementary Figure 1 for a NeuroX cluster plot of the exm27138_ver2 probe.

**Figure 1:**
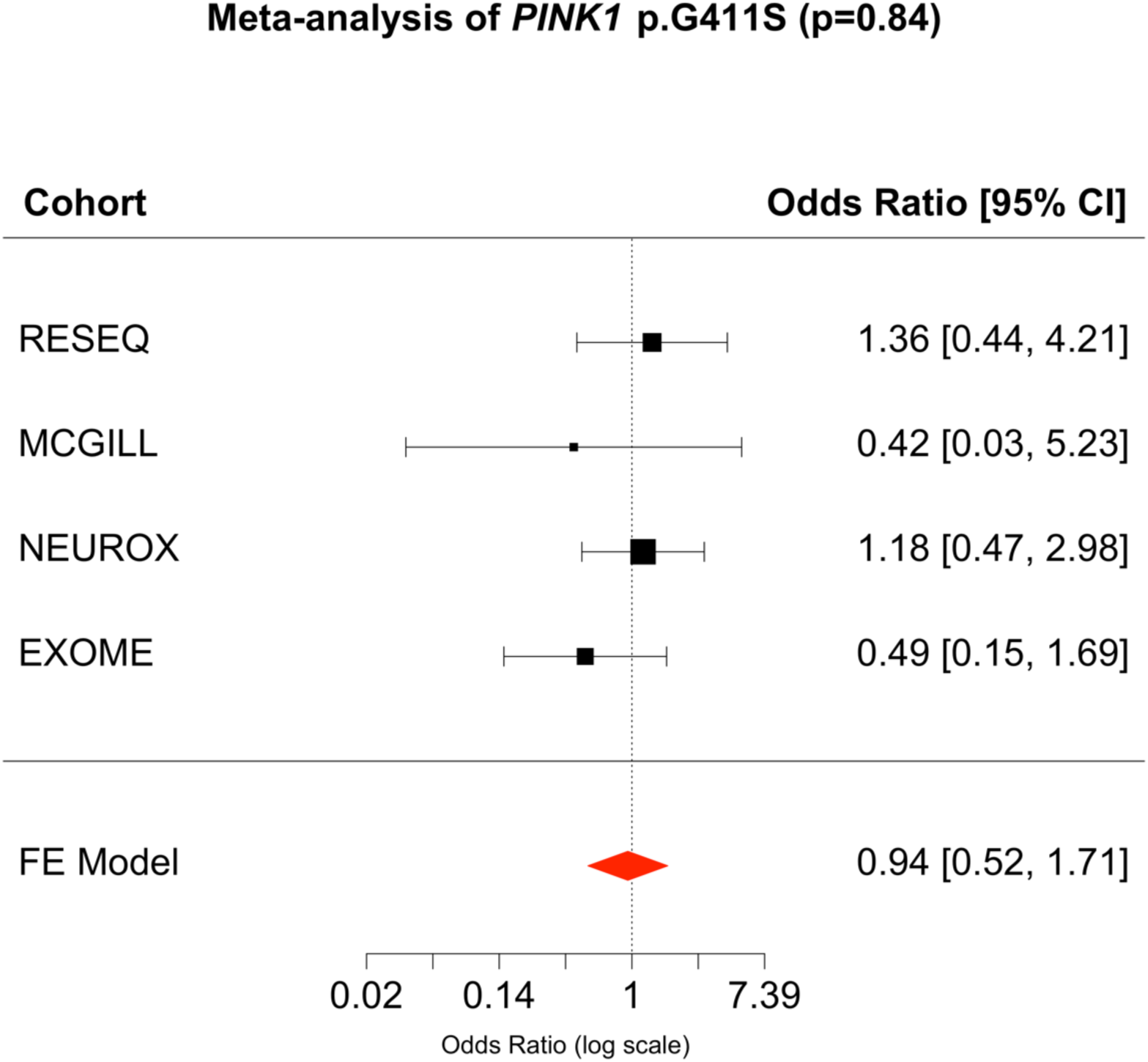
Forest plot of *PINK1* p.G411S association results per tested cohort. Sample sizes can be found in Table 1. *CI, Confidence Interval; FE, Fixed-effects*.

#### Sequencing data

Exome sequencing and resequencing data was obtained from the IPDGC totalling 5,561 cases and 5,571 controls. Data processing was performed as previously described (Blauwendraat, Kia, et al. 2018; Robak et al. 2017; Blauwendraat, Reed, et al. 2018), with variants filtered for a minimal read depth of 20, genotype quality score of greater than 20, and an alternate allele ratio of greater than 25%. If a variant was identified and the bam aligned file was available, it was visualized using the Integrative Genomics Viewer (IGV) (Thorvaldsdóttir, Robinson, and Mesirov 2013). The entire *PINK1* region (PINK1:NM_032409) was annotated using ANNOVAR (v2018-04-16) (Wang, Li, and Hakonarson 2010) and filtered for loss-of-function variants (stop-gain, frameshift insertion and frameshift deletion) and potential pathogenic variants based on ClinVar (clinvar_20190305, CLNDN = Parkinson’s disease, CLNSIG = Pathogenic). Overlapping samples were identified based on sample-ID and high PIHAT >0.8 in PLINK (v1.9). Identified duplicate samples between datasets were excluded from the larger dataset.

**Table 1:** Overview of *PINK1* p.G411S frequencies in several cohorts. Between brackets are the number of *PINK1* p.G411S variants. Data derived from Supplementary Table 1, Puschmann et al paper, and gnomAD (https://gnomad.broadinstitute.org/ accessed December 2019).

| Data-type | Cases | Controls | Cases_MAF | Controls_MAF |
| --- | --- | --- | --- | --- |
| NeuroX | 5561 (10) | 5636 (9) | 0.0009 | 0.0008 |
| Sequencing studies (IPDGC + McGill) | 6352 (14) | 6289 (24) | 0.0011 | 0.0019 |
| UKbiobank case-control (genotype + exome) | 1542 (0) | 350361 (357) | 0 | 0.00051 |
| gnomAD (European non-Finnish) | NA | 64503 (259) | NA | 0.0020 |
| gnomAD (Finnish) | NA | 12444 (247) | NA | 0.0099 |
| Puschmann et al | 2541 (19) | 2140 (5) | 0.0037 | 0.0012 |
| Puschmann other studies<sup>^</sup> | 2730 (3) | 1703 (1) | 0.00055 | 0.00029 |

The coding and regulatory regions of *PINK1* were also sequenced in a separate dataset of 1,073 cases and 961 controls from McGill University (details on this cohort were previously described (Gan-Or et al. 2019)) using Molecular Inversion Probes (MIPs) designed, targeted, and amplified as previously described (Ross et al. 2016). The MIPs library was sequenced using Illumina HiSeq 4000 platform at the McGill University and Genome Québec Innovation Centre and the full protocol is available upon request. Sequencing processing was done by Burrows-Wheeler for alignment (BWA), Genome Analysis Toolkit 29 (GATK v3.8) for post-alignment adjustments and variant calling, and ANNOVAR (Wang, Li, and Hakonarson 2010) for annotation. Quality control was performed in line with the exome sequencing data.

### UK Biobank data

#### Genotyping data

As an independent dataset we used the UKbiobank which is a population scale dataset. Imputed UKbiobank genotype data (v3) was downloaded (April 2018) (Sudlow et al. 2015; Bycroft et al. 2018). PD cases were identified using field code 42033 and controls were set as people with no report of PD, no parent with PD (data field 20107 and 20110) and age of recruitment of >60. Covariates were obtained from the data fields: genetic sex (22001) batch (22000), age of recruitment (21022), and Townsend index (189). Associations were performed using PLINK (v1.9) using a logistic regression with covariates batch, age of recruitment, Townsend index and 5 principal components to account for population stratification. Individuals were filtered for relatedness at the first cousin level (sharing proportionally more than 12.5% of alleles based on the pre-imputed data. *PINK1* p.G411S (rs45478900) was directly genotyped on the genotyping array. If a variant of interest was identified then ICD10 numbers (data field 41270) were used to identify potential movement disorders in controls.

#### Sequencing data

UKbiobank exome sequencing data (FE) was downloaded May 2019 of 49,960 individuals. The whole *PINK1* region was annotated using ANNOVAR (Wang, Li, and Hakonarson 2010) and filtered for loss-of-function variants (stop-gain, frameshift insertion and frameshift deletion) and potential pathogenic variants based on ClinVar (clinvar_20190305). The concordance rate between genotyped rs45478900 carriers and exome sequenced carriers was 100%.

### Statistical analyses and variant validation

Associations were performed using RVTESTS score test analysis (Zhan et al. 2016), followed by meta-analysis of the four cohorts using the R package metafor (Viechtbauer 2010). Additional allele frequencies were obtained from the The Genome Aggregation Database (gnomAD v2.1.1) http://gnomad.broadinstitute.org/ (Lek et al. 2016). Power calculations were performed using GAS Power Calculation (http://csg.sph.umich.edu/abecasis/cats/gas_power_calculator/) using a disease prevalence of 1% and a significance level of 0.05. In order to validate the genotyping of the *PINK1* variant, we obtained whole genome sequencing data from a parallel IPDGC project. One individual carrying the *PINK1* p.G411S variant was included in the genotyped series and heterozygous state of *PINK1* p.G411S was confirmed. Additionally, nine samples were confirmed using Sanger sequencing using previously described primers and PCR conditions for *PINK1* exon 6 PINK1_X6_F:GCTATGTCTTGCTGGTGGCTTTA and PINK1_X6_R:CAAGGCATCGAGTCTCCTGC (Brooks et al. 2009) see Supplementary Figure 2 for sequence chromatograms.

## Results

### Data overview

In order to assess the presence of *PINK1* variants in PD and control samples, we aggregated data from 5 different datasets including IPDGC data (NeuroX, Resequencing, Exome sequencing) McGill (Resequencing) and UKbiobank (Genotypes and Exome sequencing) totalling 13,708 PD cases and 362,850 controls including 376,558 individuals. In total, we identified 177 coding (missense or loss-of-function) variants in the coding region of *PINK1* (Supplemental Table 1). Out of the 177 variants, six were previously classified as pathogenic based on ClinVar and an additional 13 could be classified as pathogenic due to being clear loss of function, for a total of 19 pathogenic variants identified. An additional 11 variants were classified as of uncertain significance based on ClinVar. No individuals were identified carrying two pathogenic variants.

#### Assessment of pathogenicity of *PINK1* p.G411S

In the IPDGC and McGill datasets, we investigated 12,166 cases and 12,489 controls for their *PINK1* p.G411S status. We identified 57 heterozygous carriers, 24 cases (MAF = 0.0012) and 33 controls (MAF = 0.0015) (Table 1). Average age of assessment of controls was 60.1 (range=22-102) and average age of onset for cases was 58.3 (range=37-80). None of the controls was diagnosed with a movement disorder based on ICD10 codes. No homozygous carriers of *PINK1* p.G411S individuals were identified. These frequencies are similar to the gnomAD frequency, reporting 0.002 in Europeans. Association tests were performed for each dataset separately (NeuroX, IPDGC resequencing, IPDGC exomes, McGill) and the results of the score test analyses showed varying direction of effect across the four cohorts (Figure 1), while meta-analysis resulted in a non-significant association (OR = 0.940, 95% CI = 0.52 - 1.71, P = 0.84). Power calculations showed that there was 80% power to detect an odds ratio of 2 using this sample size (Supplementary Figure 4).

As an independent dataset we used the UKbiobank which is a population scale dataset. In total, 357 heterozygous (European) carriers of the *PINK1* p.G411S mutation were identified (average age of recruitment=56.1, range=40-70), resulting in similar frequencies as the other dataset (Table 1). Notably, only one homozygous carrier was of non-European ancestry without known neurological problems and was in their late sixties. No *PINK1* p.G411S variants were identified in PD cases (Table 1) and this variant was not included as a possibly pathogenic variant in MDSGene (www.mdsgene.org). Overall, these frequencies are highly similar to the reported gnomAD (Table 2) and again argue against pathogenicity.

#### Heterozygous pathogenic variants in cases and controls

When exploring heterozygous pathogenic variants in *PINK1* we used only sequencing data (Reseq, Exome, McGill and UKB exome) totalling 6,712 cases and 45,113 controls. Note that in the most recent PD GWAS, no signal is detected at the *PINK1* region (Supplemental Figure 3). In total, 38 individuals carried a pathogenic variant, 6 cases and 32 controls. Average age assessment of controls was 58.8 (range=40-82) and age of onset of cases was 49.5 (range=20-67). When investigating these 38 individuals further for a potential second pathogenic missense variant, assuming that this variant should be rare (MAF <0.01) no other *PINK1* coding variants were identified in the 38 individuals.

Overall percentages of pathogenic variants in cases 0.089% (6/6712) and controls 0.071% (32/45,113) indicated no clear differences. When comparing these frequencies (case/control only), a non significant P-value was identified (P=0.60, CHISQ=0.272, OR=1.26, 95% CI=0.53-3.01). Power calculations showed that there was 80% power to detect an odds ratio of 2.5 using this sample size (Supplementary Figure 5).

When assessing the frequency of loss of function variants in the gnomAD database, 23 unique variants with 46 alleles in total (excluding three gnomAD QC flagged variants) are identified in 64,599 individuals (maximal number of non-Finnish European samples in *PINK1* region) resulting in a frequency of 0.071% (under the assumption of no compound heterozygous carriers).

## Discussion

Deleterious *PINK1* variants are the second most common cause of (often young-onset) autosomal recessive PD after mutations in *PRKN* (PARK2). Several studies reported that damaging variants found in heterozygous state could also contribute to the development of PD (Criscuolo et al. 2006; Klein et al. 2007; Eggers et al. 2010; Ricciardi et al. 2014; Puschmann et al. 2017). In this study, we screened a large number of PD patients and controls for the reported pathogenic *PINK1* p.G411S variant and other pathogenic variants.

Contrary to previous reports, our results suggest that the *PINK1* p.G411S variant is likely benign, and that other *PINK1* pathogenic variants in heterozygous state are likely not associated with increased risk for PD. Our study was, however, not designed to assess possible subtle signs of parkinsonism in heterozygous mutation carriers, as previously detected (Nürnberger et al. 2015; Weissbach et al. 2017). The frequency of the p.G411S variant in PD cases in the current data is remarkably lower compared to the previous report (Puschmann et al. 2017). Noteworthy, according to the gnomAD database, the frequency of this variant is much higher in the Northern (East) European population (Estonia, Finland and Sweden) compared to the North-Western European and Southern European frequencies.

The current study has several limitations. First, here we aggregated data from multiple sources and backgrounds which can be complex to combine. We analyzed data separately when possible and if data was merged in subsequent analysis, data origin was included as a covariate to prevent potential bias. While typically rare, genotyping and sequencing errors resulting in false positive or negative variants may still occur. Second, since *PINK1*-related PD is autosomal recessive, it is possible that in some heterozygous *PINK1* pathogenic variant PD cases carry a second damaging variant is missed or that there are other (currently) unknown potential modifiers of disease. More complex potential damaging variants like deep intronic variants with splicing effects or larger structural variants are still hard to detect using standard genotyping and sequencing techniques. By using relatively stringent criteria of pathogenicity using known ClinVar pathogenic variants or variants with clear loss-of-function mechanism, this could also result in underestimation of damaging mutation frequencies. Our analyses were done under the hypothesis that if there is an effect on risk, it should be visible with the most clear pathogenic variants.

Third, although we included a large number of individuals, the number of actual variant carriers is still relatively low and therefore we have limited power to detect small risk increases (OR <2). Additiontionally, it would be possible that variant carriers being identified as control may display subtle signs of parkinsonism requiring a movement disorder examination or will develop PD at a later stage in life. However, when comparing ages between cases and controls, cases were overall younger compared to controls. Overall, when the effect size is large enough even with relatively rare variants, clear enrichments for disease can be proved, e.g. with *LRRK2* p.G2019S. Similar to the *SNCA* p.H50Q variant (Blauwendraat, Kia, et al. 2018), despite interesting functional data, the *PINK1* p.G411S variant seems to be benign in a large sample size. It is important to note that although we included a very large amount of data, we cannot exclude that heterozygous *PINK1* pathogenic variants have a minor effect on risk for PD.

In conclusion, in the current era where large-scale genotyping and large genome sequencing studies are becoming the new standard, it is crucial for variant carriers, families, clinicians and genetic counselors to know what the potential risk is for variants of interest. Based on the current data presented here, there is insufficient evidence to classify the *PINK1* p.G411S or any other pathogenic *PINK1* variants in heterozygous state as a major risk factor or causative for PD.

## Data Availability

NA

## Acknowledgments

We would like to thank all of the subjects who donated their time and biological samples to be a part of this study. We also would like to thank all members of the International Parkinson Disease Genomics Consortium (IPDGC). See for a complete overview of members, acknowledgements and funding http://pdgenetics.org/partners and Supplementary Table 2. This work was supported in part by the Intramural Research Programs of the National Institute of Neurological Disorders and Stroke (NINDS), the National Institute on Aging (NIA), and the National Institute of Environmental Health Sciences both part of the National Institutes of Health, Department of Health and Human Services; project numbers 1ZIA-NS003154, Z01-AG000949-02 and Z01-ES101986. This research has been conducted using the UK Biobank Resource under Application Number 33601. Data used in the preparation of this article were obtained from the Parkinson’s Progression Markers Initiative (PPMI) database (www.ppmi-info.org/data). For up-to-date information on the study, visit www.ppmi-info.org. PPMI, a public-private partnership, is funded by the Michael J. Fox Foundation for Parkinson’s Research and funding partners, including AbbVie, Avid, Biogen, Bristol-Myers Squibb, Covance, GE Healthcare, Genentech, GlaxoSmithKline, Lilly, Lundbeck, Merck, Meso Scale Discovery, Pfizer, Piramal, Roche, Servier, Teva, UCB, and Golub Capital. Data and biospecimens used in preparation of this manuscript were obtained from the Parkinson’s Disease Biomarkers Program (PDBP) Consortium, part of the National Institute of Neurological Disorders and Stroke at the National Institutes of Health. Investigators include: Roger Albin, Roy Alcalay, Alberto Ascherio, DuBois Bowman, Alice Chen-Plotkin, Ted Dawson, Richard Dewey, Dwight German, Xuemei Huang, Rachel Saunders-Pullman, Liana Rosenthal, Clemens Scherzer, David Vaillancourt, Vladislav Petyuk, Andy West and Jing Zhang. The PDBP Investigators have not participated in reviewing the data analysis or content of the manuscript. DNA panels from the NINDS Human Genetics Resource Center DNA and Cell Line Repository (http://ccr.coriell.org/ninds) were used in this study, as well as clinical data. The McGill cohort was financially supported by the Michael J. Fox Foundation, the Canadian Consortium on Neurodegeneration in Aging (CCNA), Parkinson Canada, and the Canada First Research Excellence Fund (CFREF), awarded to McGill University for the Healthy Brains for Healthy Lives (HBHL) program. The access to part of the participants for this research has been made possible thanks to the Quebec Parkinson’s Network (http://rpq-qpn.ca/en/). ZGO is supported by the Fonds de recherche du Québec - Santé (FRQS) Chercheurs-boursiers award, and is a Parkinson Canada New Investigator awardee. The access to part of the participants for this research has been made possible thanks to the Quebec Parkinson’s Network (http://rpq-qpn.ca/en/).

